# Within-Patient Comparison of [¹⁸F]FC303 ([¹⁸F]Florastamin) and [⁶⁸Ga]Ga-PSMA-11 PET/CT in Prostate Cancer: Protocol-Conditional Biodistribution and Quantitative Non-Interchangeability

**DOI:** 10.64898/2026.05.28.26354302

**Authors:** Whi-An Kwon, Sohyun Park, Ryul Kim, Woong Jin Lee, Chansoo Park, Tae-Sung Kim, Jae Young Joung

## Abstract

**Background:** Prostate-specific membrane antigen (PSMA) PET/CT is central to prostate cancer staging and theranostic workflows. To our knowledge, no direct within-patient comparison of [¹⁸F]FC303 ([¹⁸F]Florastamin) and [⁶⁸Ga]Ga-PSMA-11 has been reported. We performed a preliminary paired method-comparison study under non-harmonized acquisition protocols.

**Patients and Methods:** Twenty patients with histologically confirmed prostate cancer underwent [⁶⁸Ga]Ga-PSMA-11 PET/CT (185 ± 37 MBq, 60 ± 10 min) followed by [¹⁸F]FC303 PET/CT (370 ± 37 MBq, 105 ± 15 min) on the same PET/CT system within each patient (median interval, 29.5 days). Index targets were anatomically matched to the biopsied or surgically sampled lesion or target region. The primary malignant set included 18 histologically malignant targets; two histology-negative or indeterminate targets were included only in sensitivity analysis. Fixed [⁶⁸Ga]Ga-PSMA-11-first scan order and the 45-min uptake-time difference were central interpretive constraints.

**Results:** Across five predefined reference organs, [¹⁸F]FC303 showed lower SUVmean than [⁶⁸Ga]Ga-PSMA-11 (all Benjamini–Hochberg-adjusted p < 0.001; [⁶⁸Ga]/[¹⁸F]FC303 geometric mean ratio [GMR], 1.29–3.89). In the primary malignant set, [¹⁸F]FC303 lesion SUVmax was lower than [⁶⁸Ga]Ga-PSMA-11 (median, 11.3 vs 18.1; paired median difference, −5.50; 95% CI, −6.85 to −2.90; Wilcoxon p = 8.4 × 10⁻⁴), with strong rank correlation (Spearman ρ = 0.90). Passing–Bablok regression yielded β = 1.13 (95% CI, 1.04–1.45), and log-Bland–Altman GMR (FC303/[⁶⁸Ga]) was 0.75, consistent with proportional non-interchangeability. Tumor-to-liver and tumor-to-mediastinum ratios did not differ significantly (GMR, 1.17 [95% CI, 0.94–1.45] and 0.96 [0.80–1.15], respectively); the study was not powered for equivalence. The n = 20 sensitivity analysis showed consistent directionality.

**Conclusions:** Under non-harmonized acquisition conditions, [¹⁸F]FC303 showed lower physiologic reference-organ SUVmean and malignant target-region SUVmax than [⁶⁸Ga]Ga-PSMA-11, whereas tumor-to-liver and tumor-to-mediastinum ratios were not significantly different. Absolute SUVs were not interchangeable; [⁶⁸Ga]Ga-PSMA-11-derived SUV thresholds should not be directly transferred to [¹⁸F]FC303 without tracer-specific calibration.

---

Prostate-specific membrane antigen (PSMA) PET/CT is a central imaging modality in prostate cancer, with expanding roles in primary staging, restaging after biochemical recurrence, and theranostic treatment selection.^1, 2^ The joint EANM/SNMMI procedural guideline version 2.0 and contemporary EAU guideline frameworks recognize PSMA PET as the most sensitive imaging modality for detecting metastatic disease in appropriate clinical settings and emphasize tracer-specific performance characteristics.^1, 3^

[⁶⁸Ga]Ga-PSMA-11 remains the reference comparator in most PSMA-targeted PET studies.^4, 5^ Interest in ¹⁸F-labeled PSMA tracers has grown because fluorine-18 offers favorable physical imaging properties, enables centralized production and broader distribution, and supports more standardized clinical implementation.^6, 7^ A recent systematic review and meta-analysis demonstrated that several ¹⁸F-labeled PSMA tracers achieve lesion detection rates comparable to [⁶⁸Ga]Ga-PSMA-11 while retaining tracer-specific biodistribution features.^8^ Within-patient head-to-head comparisons are particularly informative, as between-cohort analyses are confounded by differences in disease burden, prior treatments, PSA levels, and interpretation criteria.^9, 10^

[¹⁸F]FC303 ([¹⁸F]Florastamin) has demonstrated promising preliminary clinical performance. In a first-in-human microdose trial, [¹⁸F]FC303 showed favorable biodistribution, progressive tumor uptake, and high tumor-to-background ratio without tracer-related adverse events.^11^ Subsequent prospective studies reported favorable tumor-to-background ratios and lesion detection in patients with suspected prostate cancer^12^ and diagnostic accuracy comparable to multiparametric MRI with higher specificity in intermediate-risk patients.^13^ To date, however, within-patient comparison data for [¹⁸F]FC303 against [⁶⁸Ga]Ga-PSMA-11 are not available, unlike the established evidence reported for other ¹⁸F-labeled PSMA tracers, including [¹⁸F]PSMA-1007, and [¹⁸F]DCFPyL, encompassing both biodistribution characterization and comparative evaluation against [⁶⁸Ga]Ga-PSMA-11.^14–17^

We conducted a paired within-patient comparison of [¹⁸F]FC303 and [⁶⁸Ga]Ga-PSMA-11 PET/CT under non-harmonized clinical acquisition protocols, focusing on normal-organ biodistribution, index target lesion uptake, and tumor-to-background contrast. Differences in uptake time and fixed scan order are central interpretive constraints rather than peripheral limitations. We distinguish lesion-level or target-region verified malignant index lesions from broader histology-sampled target lesion populations, with corresponding primary and sensitivity analyses.

## PATIENTS AND METHODS

### Study Design and Patients

This retrospective observational study was a paired within-patient comparison of [¹⁸F]FC303 and [⁶⁸Ga]Ga-PSMA-11 PET/CT, designed as an exploratory benchmarking analysis. Clinical and imaging data were retrieved from the National Cancer Center Hospital (Goyang, Republic of Korea) archive for patients evaluated between June 13, 2023 and March 26, 2025. All participants were enrolled in the parent FC303-3-2 Phase 3 multicenter prospective diagnostic accuracy trial (NCT05936658); index lesion handling is described under Index Target Lesion Selection and Histopathologic Verification.

Inclusion criteria were: (i) histologically confirmed prostate cancer; (ii) both PET/CT examinations within 90 days; (iii) complete imaging datasets suitable for quantitative analysis; and (iv) no documented initiation, discontinuation, or switch of systemic anticancer therapy between examinations based on electronic medical records and parent-trial records. Exclusion criteria were insufficient image quality, incomplete imaging protocols, or missing essential eligibility data. Twenty patients met these criteria. The median interval between PET/CTs was 29.5 days (IQR 25.8–36.0); [⁶⁸Ga]Ga-PSMA-11 preceded [¹⁸F]FC303 in all patients. The retrospective database did not systematically capture paired PSA values at the time of [¹⁸F]FC303 PET/CT nor detailed on-treatment status (ongoing ADT duration or ARPI exposure) beyond what is summarized in Table 1; this is acknowledged as a limitation.

**Table 1.**
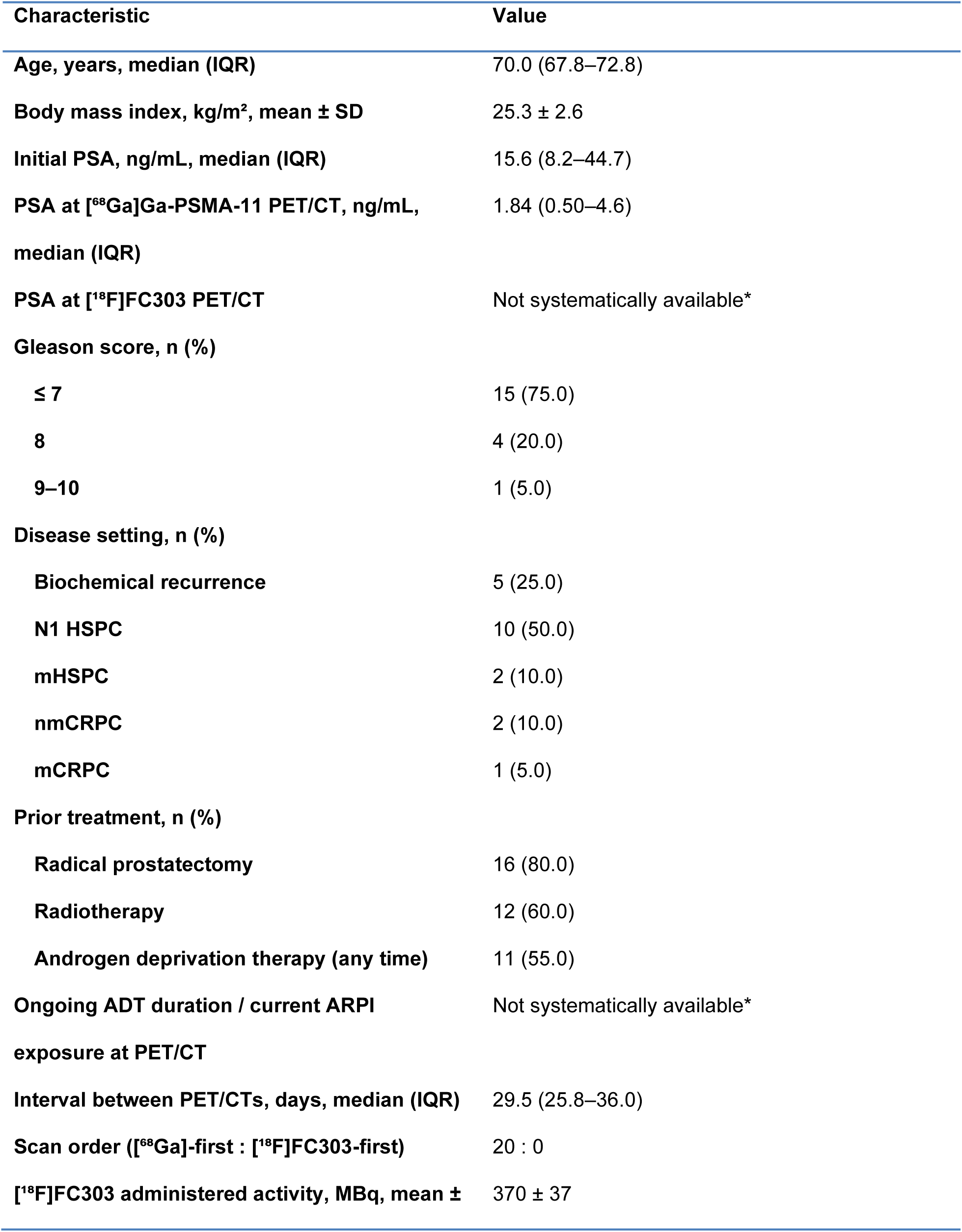
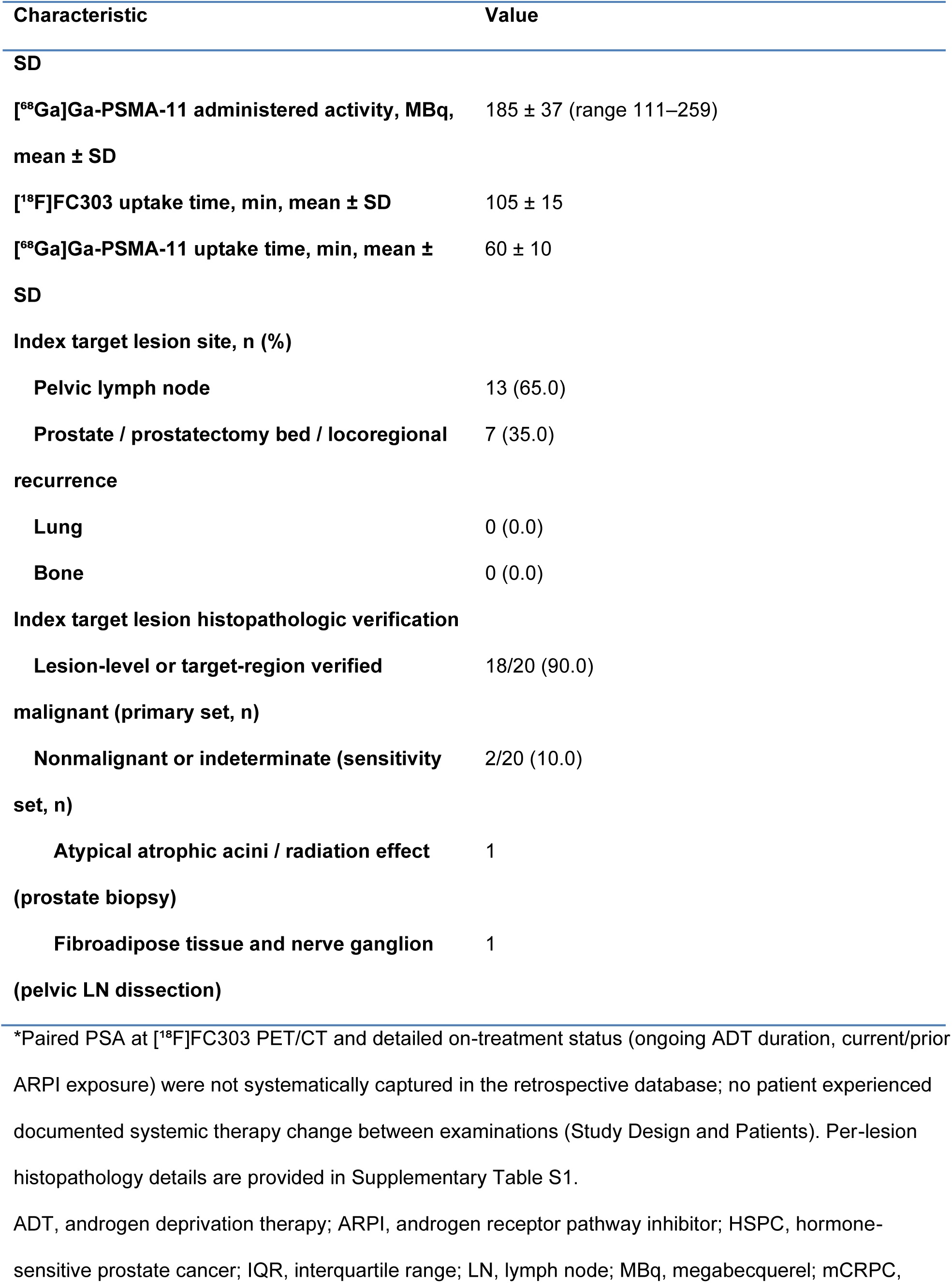
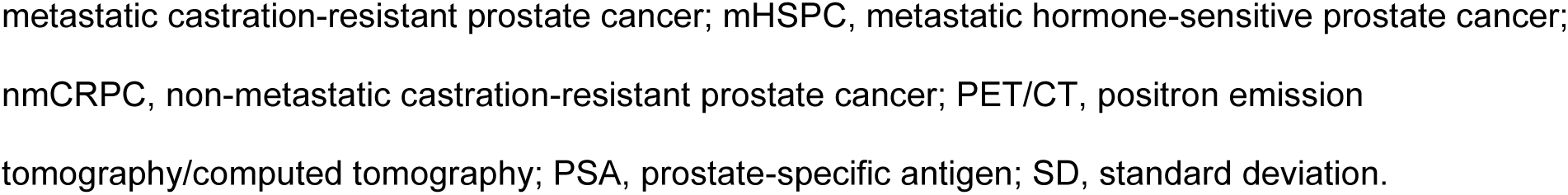
Patient, Imaging, and Index Target Lesion Characteristics (n = 20 patients)

The study was approved by the Institutional Review Board of the National Cancer Center of Korea (IRB No. NCC2023-0090). Written informed consent was waived given the retrospective analysis of previously consented trial participants, and all procedures complied with the Declaration of Helsinki.^18^

### Radiopharmaceutical Preparation

[¹⁸F]FC303 was manufactured under Good Manufacturing Practice conditions at the FutureChem production facility (automated synthesis module; HPLC purification; radiochemical purity > 95%). [⁶⁸Ga]Ga-PSMA-11 was synthesized using a ⁶⁸Ge/⁶⁸Ga generator (BIK Therapeutics, Republic of Korea) per European Pharmacopoeia guidelines (radiochemical yield > 70%, radiochemical purity > 95%).

### PET/CT Acquisition

Whole-body PET/CT from vertex to mid-thigh was performed in 3D mode on one of two digital PET/CT systems (Discovery MI or Omni Legend; GE Healthcare, USA) with a 500-mm field of view. Acquisition time per bed position was 2 min 30 s (Discovery MI) or 3 min 30 s (Omni Legend). Low-dose CT was acquired for attenuation correction and anatomic localization (120 kVp, tube current modulation, 2.5–3.75 mm slice thickness). PET reconstruction followed scanner-specific vendor-recommended protocols per the institutional nuclear medicine imaging protocol (Discovery MI and Omni Legend; 5.0-mm Gaussian post-filter, 256 × 256 matrix). Each patient’s paired examinations were acquired on the same system.

[⁶⁸Ga]Ga-PSMA-11 was administered intravenously at 3.0–3.7 MBq/kg (mean 185 ± 37 MBq), with imaging at 60 ± 10 min post-injection. [¹⁸F]FC303 was administered at 370 ± 37 MBq with imaging at 105 ± 15 min post-injection. These times reflect the short physical half-life of gallium-68 and the favorable pharmacokinetics of [¹⁸F]FC303 at delayed timepoints. The 45-min uptake-time difference, the 2-fold activity difference, and the fixed [⁶⁸Ga]Ga-PSMA-11-first scan order constitute central interpretive constraints addressed in the Discussion.

### Image Interpretation and Volume-of-Interest Methodology

PET images were independently reviewed by two board-certified nuclear medicine physicians with more than 10 years of experience in PSMA PET imaging. Readers were blinded to clinical and pathologic information and to the paired alternate-tracer result; tracer identity could not be fully masked because whole-body biodistribution patterns, acquisition parameters, and urinary activity patterns inherently reveal the tracer. Final lesion classifications were obtained by consensus read; formal inter-reader agreement statistics for the independent pre-consensus reads are not reported because pre-consensus classifications were not retained in the retrospective database. Lesions were defined as focal tracer uptake exceeding the surrounding background and inconsistent with physiologic uptake patterns, per the EANM/SNMMI guideline.^1^

For normal-organ biodistribution, 1-cm³ spherical volumes of interest (VOIs) were placed in lesion-free, artifact-free regions using standardized anatomical rules: liver right lobe (segment VII/VIII), spleen mid-body, mediastinal blood pool (ascending aorta at the level of the main pulmonary artery), bilateral parotid glands (averaged), and bilateral renal cortex mid-polar regions (averaged). SUVmean was recorded for each organ. The same anatomical VOI positions were used on both tracer examinations for each patient.

For each patient, the protocol-designated index target lesion (see Index Target Lesion Selection and Histopathologic Verification) was analyzed. A VOI was placed to encompass the lesion on each tracer examination and SUVmax was recorded. For index targets that were visually non-detected on one tracer, the volume of interest was positioned within the anatomically matched, protocol-designated target region defined by CT anatomy, the procedural target description, and the paired-tracer and pathology reference, and the resulting value was interpreted as target-region SUVmax rather than detected-lesion SUVmax; visual detection classification was adjudicated separately from quantitative VOI placement. Tumor-to-liver (T/L = lesion SUVmax / liver SUVmean) and tumor-to-mediastinum (T/M = lesion SUVmax / mediastinal blood-pool SUVmean) ratios were calculated. Formal urinary bladder and ureteral SUV quantification was not pre-specified and was not feasible to retrospectively re-measure; this is acknowledged as a limitation given that all analyzed index target lesions were locoregional pelvic or prostate/prostate-bed targets.

### Index Target Lesion Selection and Histopathologic Verification

All patients were derived from the FC303-3-2 Phase 3 diagnostic accuracy trial. In the parent trial, biopsy-accessible target lesions were prospectively identified on baseline conventional imaging within 60 days before [¹⁸F]FC303 PET/CT and underwent image-guided biopsy or surgical resection within 12 h to 28 days after [¹⁸F]FC303 PET/CT. Local pathologists were blinded to PET/CT findings.

For the present study, one protocol-designated target lesion per patient was selected as the index target lesion; selection was not based on uptake intensity on either tracer. Lesion matching between PET-identified uptake and the histology-sampled target was adjudicated using laterality, anatomical compartment, procedural target description in the surgical/biopsy report, and pathology report concordance. For pelvic lymph node dissection specimens, pathology was reported at the nodal basin or regional level (e.g., “x of y nodes metastatic”) rather than by individual PET-imaged node; such cases are treated as lesion-level or target-region histopathology.

Two analysis sets were defined based on the granularity of histopathologic verification at the analyzed site:

(i) Primary malignant set (n = 18): histologically verified malignant at the analyzed anatomical site, comprising pelvic lymph node metastases (lesion-level or nodal basin verification) and prostate / prostatectomy-bed / perirectal adenocarcinoma recurrence (lesion-level verification).

(ii) Sensitivity set (n = 20): primary set plus two histology-negative or indeterminate index target lesions (one prostate biopsy showing atypical atrophic acini suspicious for radiation therapy effect; one pelvic lymph node dissection showing fibroadipose tissue and nerve ganglion without evidence of tumor).

Normal-organ biodistribution analyses were performed in all 20 patients because they are patient-level rather than lesion-dependent. Descriptive index target lesion classification concordance was evaluated for the single protocol-designated index target lesion in each of the 20 patients. Per-patient histopathology details are provided in Supplementary Table S1.

### Index Target Lesion Classification

As a descriptive secondary analysis, the single protocol-designated index target lesion in each patient was visually categorized on each tracer examination as detected, equivocal, or non-detected. Because a uniform lesion-level truth standard was not applied across all anatomical contexts and the sample size was not designed to establish diagnostic equivalence, this analysis is reported as descriptive classification concordance rather than as a comparative diagnostic accuracy endpoint.

### Statistical Analysis

SUV-derived variables are reported as median (interquartile range) because of their skewed distributions; demographic and acquisition variables are reported as median (interquartile range) or mean ± SD, as appropriate. Companion mean ± SD values for the primary quantitative endpoints are provided in Supplementary Tables S2A and S2B. The primary endpoint for the present exploratory analysis was the paired median difference in malignant index lesion SUVmax (primary set, n = 18) between the two tracers, tested using the Wilcoxon signed-rank test with bootstrap 95% confidence intervals (B = 10,000). For normal-organ biodistribution (n = 20), paired Wilcoxon signed-rank tests were applied across five predefined organs with Benjamini–Hochberg false discovery rate adjustment.^19^ Fold differences between tracers were reported as geometric mean ratios (GMR) with 95% CI based on log-transformed paired ratios.

Between-tracer association for malignant index lesion SUVmax was assessed using Spearman correlation (with Pearson and log-Pearson as sensitivity analyses, reported in Supplementary Material), whereas quantitative agreement and interchangeability were assessed using Bland–Altman analysis on both raw and log-transformed scales reporting bias with 95% CI and 95% limits of agreement,^20^ and Passing–Bablok regression ^21^ modeled as [⁶⁸Ga]Ga-PSMA-11 SUVmax = α + β × [¹⁸F]FC303 SUVmax; Deming regression with λ = 1 is reported for consistency. The Hodges–Lehmann estimator ^22^ was computed as a paired effect-size measure. It is related but not identical to the paired median difference (the Hodges–Lehmann estimator is the median of all pairwise Walsh averages of differences), and small numerical discrepancies are expected in moderately skewed paired data.

For tumor-to-liver and tumor-to-mediastinum ratios, we report paired log-ratio geometric mean ratios ([¹⁸F]FC303/[⁶⁸Ga]Ga-PSMA-11) with 95% CI, paired median differences with bootstrap 95% CI, and Wilcoxon p values. The study was not powered for equivalence or non-inferiority; 95% CIs are intended to inform interpretation rather than support equivalence claims. Exploratory lesion-site analyses were summarized descriptively; for the pelvic lymph-node subgroup, the paired SUVmax difference was additionally assessed using a two-sided exact Wilcoxon signed-rank test where feasible, with zero-difference pairs excluded according to the Wilcoxon convention. All quantitative analyses were repeated in the n = 20 sensitivity set. A two-sided p < 0.05 was considered statistically significant. Analyses were performed in Python 3.11 (SciPy 1.11, statsmodels 0.14) and GraphPad Prism 9.0 (GraphPad Software, San Diego, CA). Paired bootstrap confidence intervals were percentile intervals generated by patient-level resampling of paired observations (B = 10,000), and Wilcoxon signed-rank tests used the standard convention of excluding zero-difference pairs. For directional clarity, geometric mean ratios for normal-organ and lesion SUVmax are expressed as [⁶⁸Ga]Ga-PSMA-11/[¹⁸F]FC303 (values > 1 denote lower measured [¹⁸F]FC303 uptake), whereas geometric mean ratios for tumor-to-background ratios and log-transformed Bland–Altman analyses are expressed as [¹⁸F]FC303/[⁶⁸Ga]Ga-PSMA-11.

## Results

### Patient Characteristics

Twenty patients met the eligibility criteria (Table 1). Median age was 70.0 years (IQR 67.8–72.8) and mean body mass index 25.3 ± 2.6 kg/m². Initial PSA was 15.6 ng/mL (IQR 8.2–44.7), and PSA at the time of [⁶⁸Ga]Ga-PSMA-11 PET/CT was 1.84 ng/mL (IQR 0.50–4.6). Paired PSA values at the time of [¹⁸F]FC303 PET/CT were not systematically available in the retrospective database. Gleason score was ≤ 7 in 15 patients (75%), 8 in 4 (20%), and 9–10 in 1 (5%). Disease settings comprised biochemical recurrence (n = 5; 25%), N1 hormone-sensitive prostate cancer (N1 HSPC; n = 10; 50%), metastatic HSPC (n = 2; 10%), non-metastatic castration-resistant prostate cancer (nmCRPC; n = 2; 10%), and metastatic CRPC (n = 1; 5%). Androgen deprivation therapy (ADT) had been administered at any time in 11 patients (55%); detailed ongoing ADT duration and prior/current androgen receptor pathway inhibitor (ARPI) exposure were not retrievable from the retrospective database. The median interval between examinations was 29.5 days (IQR 25.8–36.0); [⁶⁸Ga]Ga-PSMA-11 PET/CT preceded [¹⁸F]FC303 PET/CT in all patients.

### Index Target Lesion Histopathologic Verification

Histopathologic verification was available for all 20 protocol-designated index target lesions (Table 1; Supplementary Table S1). Eighteen lesions (90%) had histopathologic verification at the analyzed anatomical site and constituted the primary malignant analysis set, comprising pelvic lymph node metastases (n = 12) and prostate/prostatectomy-bed or locoregional recurrence lesions (n = 6). Two index target lesions (10%) were nonmalignant or indeterminate on histopathology—one prostate needle biopsy showing atypical atrophic acini suspicious for radiation therapy effect, and one right pelvic lymph node dissection showing fibroadipose tissue and nerve ganglion—and were evaluated in the n = 20 sensitivity set. Across all 20 protocol-designated index target lesions, analyzed anatomical sites were pelvic lymph node (n = 13) and prostate/prostate-bed or locoregional recurrence (n = 7); no lung or bone index target lesion was included in the quantitative analysis.

### Normal-Organ Biodistribution (n = 20)

Across all five predefined organs, [¹⁸F]FC303 showed lower measured SUVmean than [⁶⁸Ga]Ga-PSMA-11 (all Benjamini–Hochberg-adjusted p < 0.001, Table 2). Geometric mean ratios ([⁶⁸Ga]/[¹⁸F]FC303) were spleen 3.89 (95% CI 3.48–4.34), kidneys 2.89 (2.32–3.60), parotid 2.33 (2.07–2.63), liver 1.55 (1.36–1.77), and mediastinal blood pool 1.29 (1.19–1.41). The mediastinal blood pool showed a small absolute difference (median 1.25 vs 1.50) despite statistical significance, reflecting the narrow dynamic range of this compartment. Representative paired images are shown in Figure 1. Paired within-patient normal-organ distributions are summarized in Figure 2.

**Figure 1.**
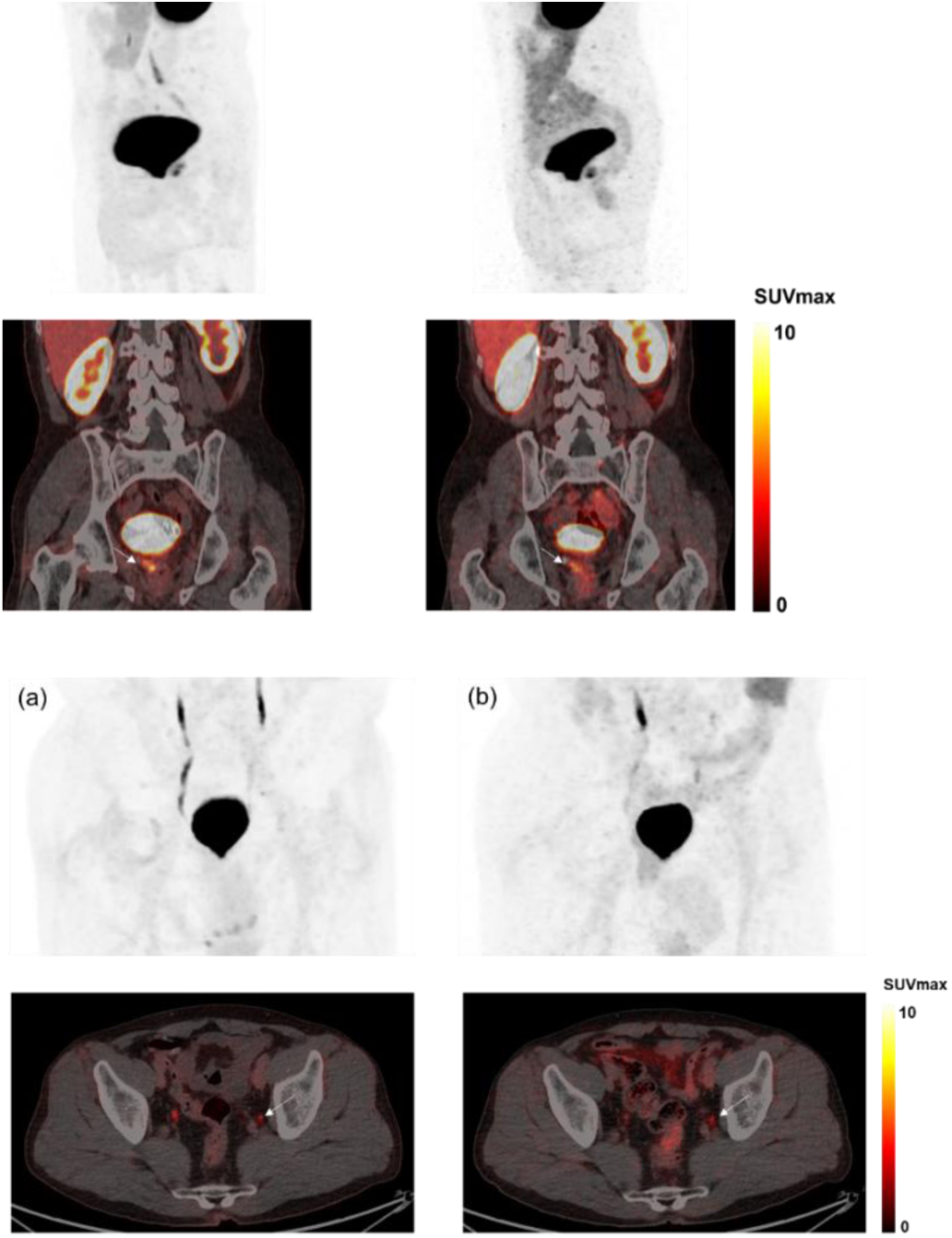
Representative paired PET/CT images. (A) Maximum intensity projection and coronal fused PET/CT images of [¹⁸F]FC303 (left) and [⁶⁸Ga]Ga-PSMA-11 (right) in a patient with suspected perirectal recurrence; the lesion demonstrates well-defined PSMA uptake (arrow) with a slightly higher lesion-to-background ratio on [¹⁸F]FC303 (LBR 7.1 vs 6.5). (B) Maximum intensity projection and axial fused PET/CT images of [¹⁸F]FC303 and [⁶⁸Ga]Ga-PSMA-11 in a patient with suspected recurrence in the left obturator region; the target lesion is visualized on both examinations (white arrow; red arrow on maximum intensity projection), with LBR 3.2 on [¹⁸F]FC303 and 2.7 on [⁶⁸Ga]Ga-PSMA-11. LBR was defined as lesion SUVmax divided by peri-lesional normal-tissue SUVmax on the same tracer examination. The lesion-to-background ratios shown in these representative cases are illustrative and exploratory only; because urinary bladder and ureteral activity were not quantitatively assessed, these examples should not be interpreted as evidence of a generalized pelvic conspicuity advantage.

**Figure 2.**
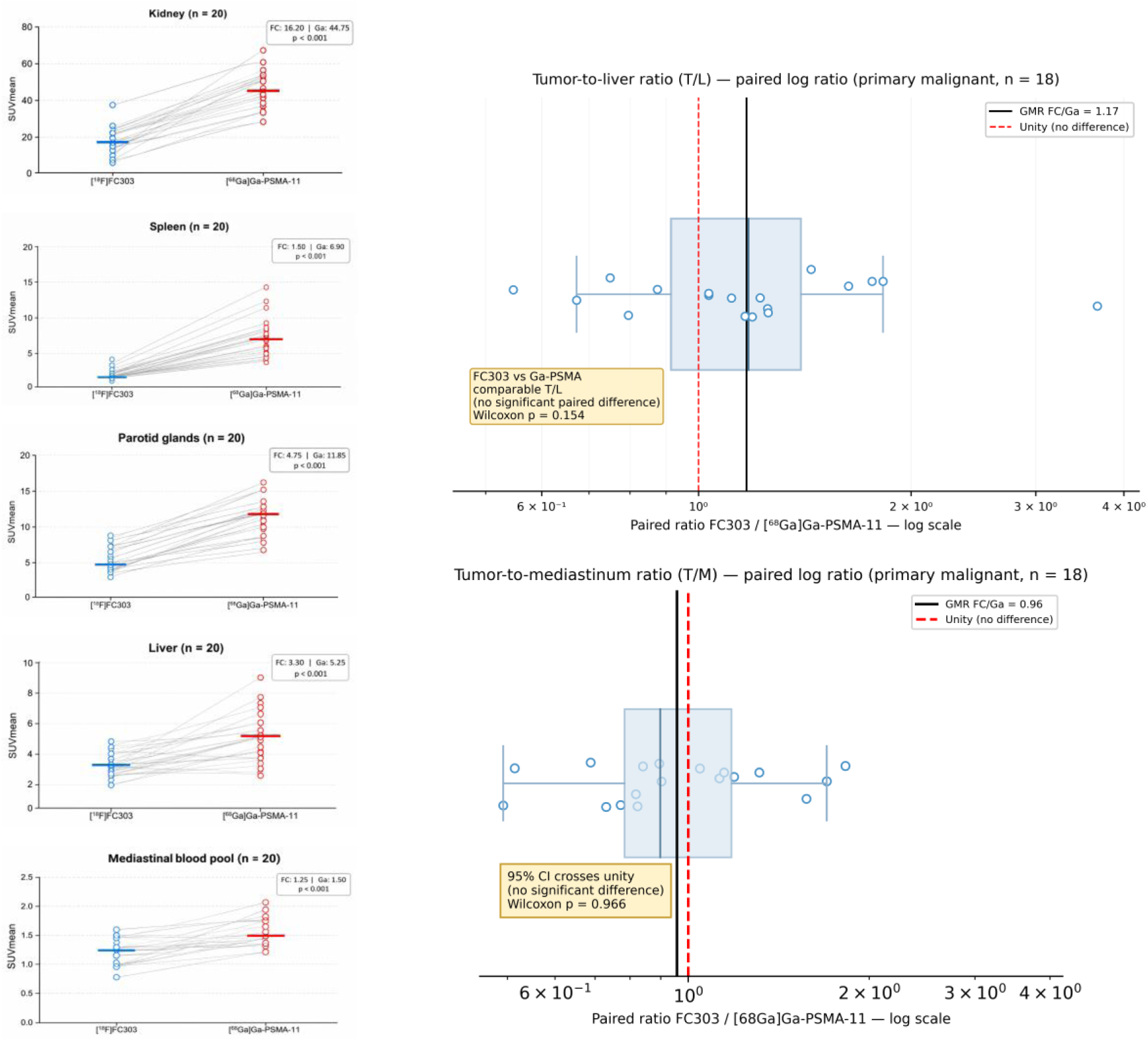
Within-patient normal-organ biodistribution and tumor-to-background ratio comparison. Left panels show paired SUVmean values for kidneys, spleen, parotid glands, liver, and mediastinal blood pool in all 20 patients. Each line connects paired measurements from the same patient. Right panels show paired log-ratio distributions for tumor-to-liver and tumor-to-mediastinum ratios in the primary malignant analysis set (n = 18), expressed as [¹⁸F]FC303/[⁶⁸Ga]Ga-PSMA-11. Normal-organ comparisons used paired Wilcoxon signed-rank tests with Benjamini–Hochberg adjustment across the five predefined organs. Tumor-to-background ratios are shown on the ratio scale; unity indicates no paired difference.

**Table 2.**
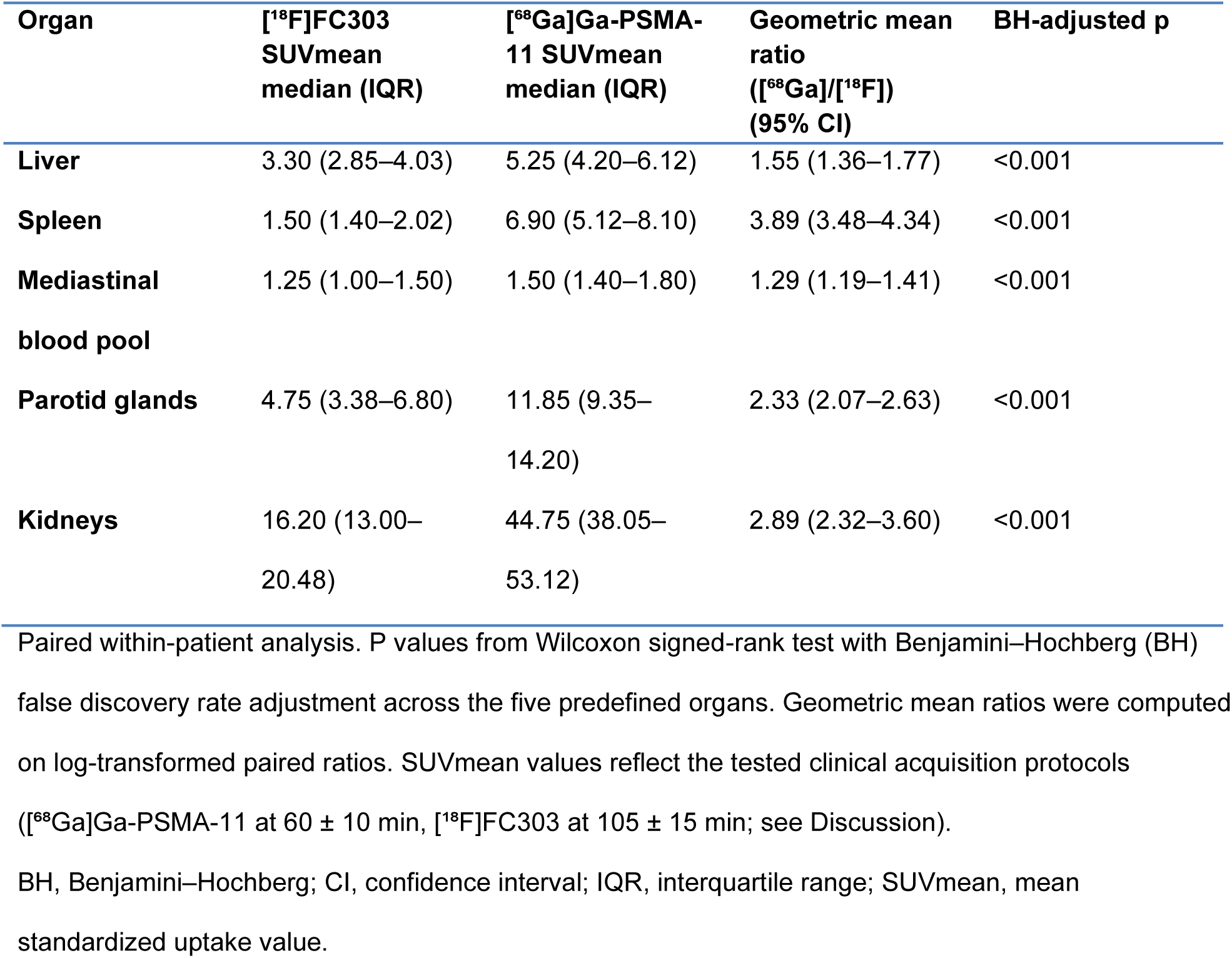
Normal-Organ Biodistribution (n = 20 patients)

### Malignant Index Target Lesion SUVmax (Primary Analysis; n = 18)

In the 18 malignant index target lesions with lesion-level or target-region histopathologic verification, SUVmax was lower with [¹⁸F]FC303 (median 11.3 [IQR 8.0–20.2]) than with [⁶⁸Ga]Ga-PSMA-11 (18.1 [10.2–29.8]). The paired median difference was −5.50 (95% bootstrap CI −6.85 to −2.90; Wilcoxon signed-rank p = 8.4 × 10⁻⁴). The Hodges–Lehmann estimator was −4.80 (95% CI −6.90 to −2.90). The paired GMR ([⁶⁸Ga]/[¹⁸F]FC303) was 1.33 (95% CI 1.15–1.54) (Table 3). The paired between-tracer correlation is shown in Figure 3.

**Figure 3.**
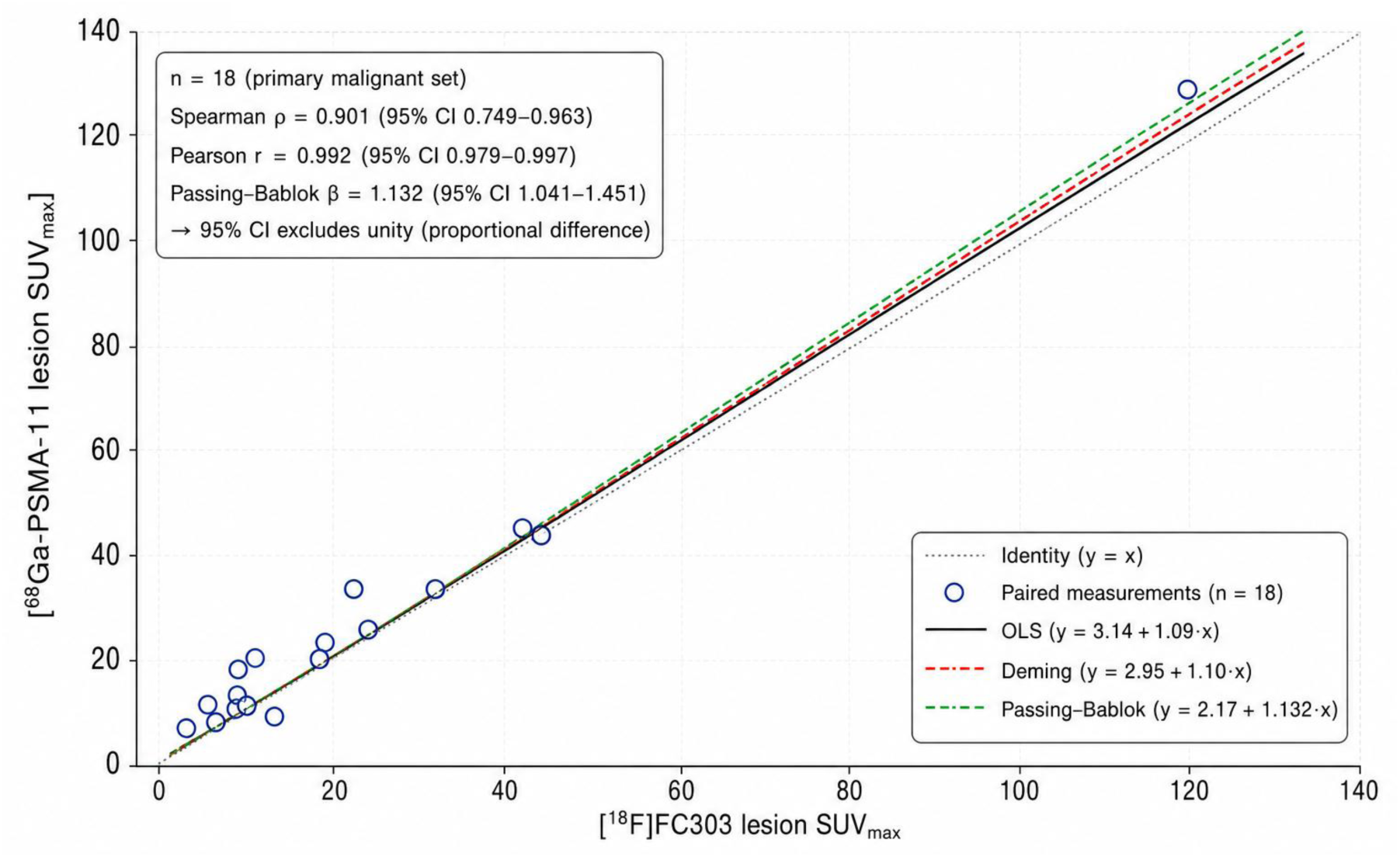
Paired malignant index target lesion SUVmax correlation. Each point represents one histologically verified malignant (lesion-level or target-region) index target lesion in the primary analysis set (n = 18). Regression models are oriented as [⁶⁸Ga]Ga-PSMA-11 SUVmax = α + β × [¹⁸F]FC303 SUVmax. The identity line indicates quantitative interchangeability. The Passing–Bablok slope 95% CI excludes unity, supporting proportional non-interchangeability despite strong rank-order concordance.

**Table 3.** Paired Index Target Lesion Metrics Across Analysis Sets.

| **Metric** | **Primary malignant**<br>**(n = 18)**<br>**histologically verified**<br>**malignant** | **Sensitivity**<br>**(n = 20)**<br>**+ nonmalignant /**<br>**indeterminate** |
| --- | --- | --- |
| **Lesion SUVmax, [<sup>18</sup>F]FC303,**<br>**median (IQR)** | 11.3 (8.0–20.2) | 11.3 (7.9–22.3) |
| **Lesion SUVmax, [<sup>68</sup>Ga]Ga-**<br>**PSMA-11, median (IQR)** | 18.1 (10.2–29.8) | 18.1 (9.9–31.3) |
| **Paired median difference (95%**<br>**CI)** | –5.50 (–6.85 to –2.90) | –5.50 (–6.95 to –2.90) |
| **Wilcoxon p** | $8.4 \times 10^{-4}$ | $3.9 \times 10^{-4}$ |
| **Hodges–Lehmann (95% CI)\*** | –4.80 (–6.90 to –2.90) | –5.32 (–7.65 to –2.90) |
| **GMR [<sup>68</sup>Ga]/[<sup>18</sup>F] (95% CI)** | 1.33 (1.15–1.54) | 1.37 (1.19–1.58) |
| **T/L paired log-ratio GMR**<br>**FC/Ga (95% CI); Wilcoxon p** | 1.17 (0.94–1.45); p = 0.15 | 1.13 (0.92–1.38); p = 0.31 |
| **T/M paired log-ratio GMR**<br>**FC/Ga (95% CI); Wilcoxon p** | 0.96 (0.80–1.15); p = 0.97 | 0.94 (0.80–1.11); p = 0.78 |

T/L = tumor-to-liver ratio = lesion SUVmax / liver SUVmean; T/M = tumor-to-mediastinum ratio = lesion SUVmax / mediastinal blood-pool SUVmean. Paired log-ratio GMRs are expressed as [<sup>18</sup>F]FC303/[<sup>68</sup>Ga]Ga-PSMA-11. The study was not powered for equivalence or non-inferiority; 95% CIs crossing unity for T/L and T/M indicate absence of detected difference rather than evidence of equivalent contrast. \*The paired median difference and Hodges–Lehmann estimator are related but distinct: the paired median difference is the median of within-patient differences, whereas the Hodges–Lehmann estimator is the median of pairwise Walsh averages of the differences; small numerical discrepancies are expected in moderately skewed paired data.
CI, confidence interval; FC, [<sup>18</sup>F]FC303; Ga, [<sup>68</sup>Ga]Ga-PSMA-11; GMR, geometric mean ratio; IQR, interquartile range; SUVmean, mean standardized uptake value; SUVmax, maximum standardized uptake value.

### Tumor-to-Background Ratios (Primary Analysis; n = 18)

For the primary malignant set (n = 18), tumor-to-liver ratio was 4.09 (IQR 2.55–6.05) for [¹⁸F]FC303 and 3.11 (IQR 1.70–5.17) for [⁶⁸Ga]Ga-PSMA-11 (paired log-ratio GMR FC303/[⁶⁸Ga] = 1.17, 95% CI 0.94–1.45; Wilcoxon p = 0.15). Tumor-to-mediastinum ratio was 10.70 (IQR 6.02–19.90) and 12.03 (IQR 6.36–20.48), respectively (paired log-ratio GMR = 0.96, 95% CI 0.80–1.15; Wilcoxon p = 0.97). Neither comparison reached statistical significance, and the 95% CIs for both paired log ratios crossed unity; however, the study was not powered for equivalence or non-inferiority, and these results should be interpreted as absence of detected difference rather than evidence of equivalent contrast. The directionally higher T/L ratio and similar T/M ratio with [¹⁸F]FC303 are compatible with the lower physiologic background observed in normal-organ analyses. Paired tumor-to-background log-ratio distributions are shown in Figure 2 (right panels). Peri-lesional lesion-to-background ratios were assessed only as an exploratory, hypothesis-generating endpoint outside the prespecified endpoint hierarchy and are reported in Supplementary Table S3.

### Method Comparison and Agreement (Primary Analysis; n = 18)

Paired malignant index target lesion SUVmax values showed strong rank-order and log-scale concordance between tracers (Spearman ρ = 0.901, 95% CI 0.749–0.963; log-transformed Pearson r = 0.946, 95% CI 0.859–0.980). The raw Pearson correlation was higher (r = 0.992, 95% CI 0.979–0.997) but was disproportionately influenced by a single high-range observation; it is therefore reported only as a supporting metric, with the rank-order and log-scale estimates—and the leave-one-out sensitivity analysis (Supplementary Material)—taken as the primary association metrics, whereas Bland–Altman analysis and regression-based method comparison (below) were used to assess quantitative agreement and interchangeability. Raw Bland–Altman analysis yielded a mean bias of −4.93 (95% CI −7.06 to −2.80; 95% limits of agreement −13.31 to +3.46). Log-transformed (geometric) Bland–Altman analysis showed a geometric mean ratio ([¹⁸F]FC303/[⁶⁸Ga]Ga-PSMA-11) of 0.75, indicating that [¹⁸F]FC303 SUVmax was approximately 25% lower than paired [⁶⁸Ga]Ga-PSMA-11 SUVmax on the geometric scale, with 95% limits of agreement of 0.42× to 1.35× on the ratio scale.

Passing–Bablok regression (modeled as [⁶⁸Ga]Ga-PSMA-11 SUVmax = α + β × [¹⁸F]FC303 SUVmax) yielded β = 1.132 (95% CI 1.041–1.451) and α = 2.17; the slope 95% CI excluded unity, consistent with proportional differences across the measurement range in addition to the systematic negative offset. Deming regression (λ = 1) gave a consistent point estimate (slope 1.10; intercept 2.95). Table 4 and Figure 4 summarize these association and method-comparison analyses. Expanded correlation and regression results, including leave-one-out sensitivity and Cook’s distance, are provided in the Supplementary Material. Raw and log-transformed Bland–Altman analyses and paired SUVmax scatter plots for the primary and sensitivity sets are shown in Supplementary Figures S1 and S2.

**Figure 4.**
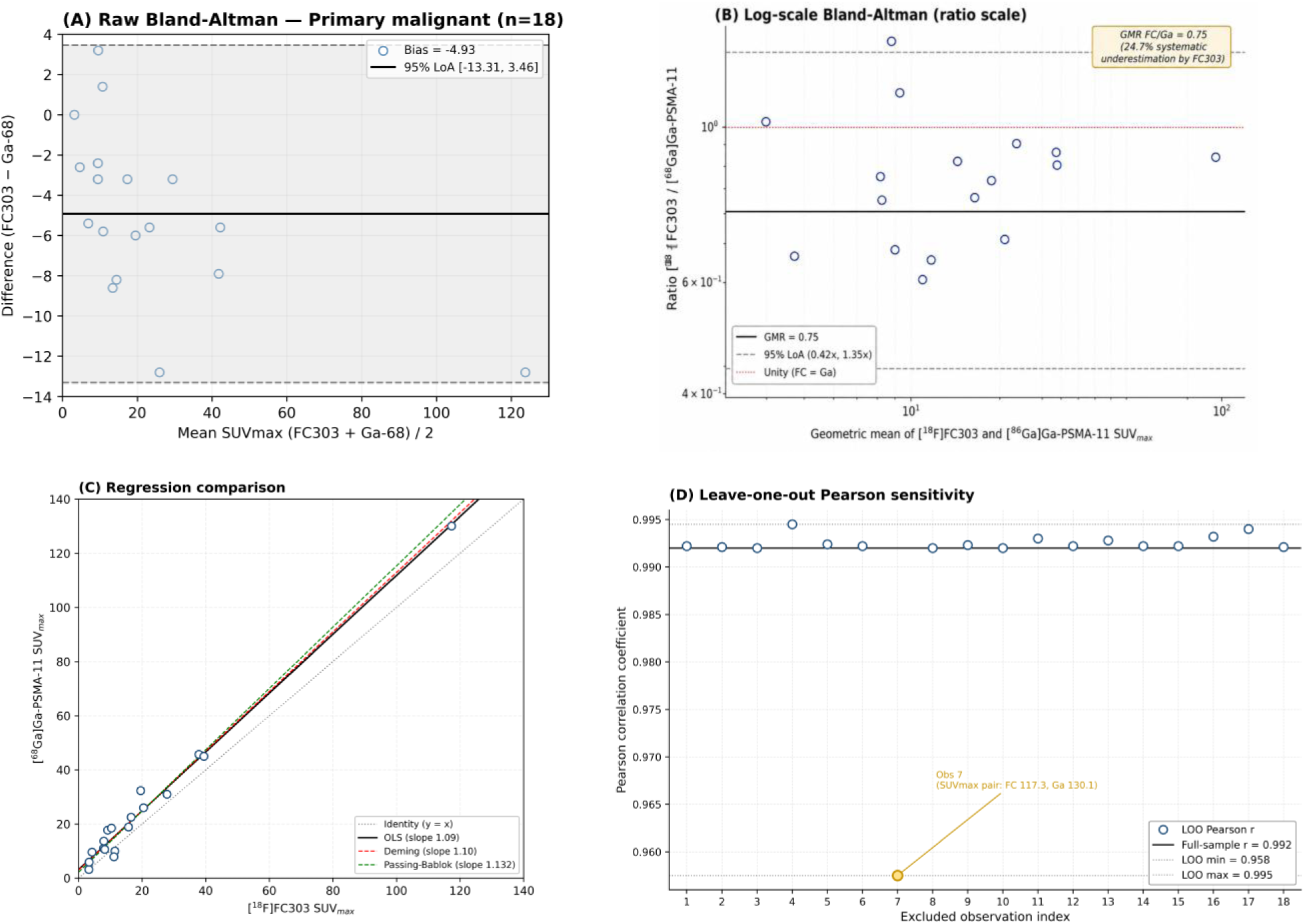
Method-comparison analyses for malignant index target lesion SUVmax. (A) Raw Bland–Altman analysis. (B) Log-transformed Bland–Altman analysis on the ratio scale. (C) Regression comparison with identity reference. (D) Leave-one-out Pearson correlation sensitivity analysis. Bias and limits of agreement are reported for [¹⁸F]FC303 relative to [⁶⁸Ga]Ga-PSMA-11; regression orientation is [⁶⁸Ga]Ga-PSMA-11 as the dependent variable.

**Table 4.** Association and Method-Comparison Analyses for Malignant Index Target Lesion SUVmax (Primary Malignant Set, n = 18; Sensitivity Analysis in Supplementary Table S5)

| Analysis | Primary malignant (n = 18) | Interpretation |
| --- | --- | --- |
| **Spearman $\rho$ (95% CI)** | 0.901 (0.749–0.963) | Robust rank concordance |
| **Pearson $r$ (95% CI)** | 0.992 (0.979–0.997) | Strong; leverage assessed in Supplementary Material |
| **Bland–Altman raw bias (95% CI)** | –4.93 (–7.06 to –2.80) | Systematic offset |
| **Bland–Altman raw 95% LoA** | –13.31 to +3.46 |  |
| **Bland–Altman log (geometric), GMR FC/Ga** | 0.75 (≈25% lower, geometric) | Consistent with raw BA |
| **Log-BA 95% LoA (ratio scale)** | 0.42× to 1.35× | |
| **Passing–Bablok slope $\beta$ (95% CI)†** | 1.132 (1.041–1.451) | CI excludes 1.0; proportional differences |
| **Passing–Bablok intercept $\alpha$†** | 2.17 |  |
| **Deming slope ($\lambda = 1$)†** | 1.10 | Consistent with Passing–Bablok |

†Passing–Bablok and Deming regressions were modeled as [ $^{68}\text{Ga}$ ]Ga-PSMA-11 SUVmax = $\alpha$ + $\beta$ × [ $^{18}\text{F}$ ]FC303 SUVmax; slope > 1 indicates that [ $^{68}\text{Ga}$ ]Ga-PSMA-11 scales up more per unit [ $^{18}\text{F}$ ]FC303. BA, Bland–Altman; CI, confidence interval; FC, [ $^{18}\text{F}$ ]FC303; Ga, [ $^{68}\text{Ga}$ ]Ga-PSMA-11; GMR, geometric mean ratio; LoA, limits of agreement; SUVmax, maximum standardized uptake value; $\alpha$ , regression intercept; $\beta$ , regression slope; $\lambda$ , error-variance ratio (Deming regression).

### Index Target Lesion Classification Concordance (n = 20)

Across all 20 histology-sampled index target lesions, overall descriptive concordance of visual classification was 75.0% (15/20; Cohen’s kappa 0.48; 95% CI provided in Supplementary Table S4). For the reported binary concordance analysis, equivocal and non-detected final consensus calls were collapsed as not detected. Binary consensus classification differed in 5 of 20 protocol-designated target regions; per-tracer detection counts are reported in Supplementary Table S4. This analysis was neither powered nor designed to compare diagnostic detection performance between tracers. Notably, [¹⁸F]FC303 was scored as detected in numerically more index targets than [⁶⁸Ga]Ga-PSMA-11 (14/20 vs 11/20) despite its lower SUVmax; given the later [¹⁸F]FC303 acquisition time and lower physiologic background, this numerical difference should not be interpreted as detection superiority of either tracer. Because no uniform lesion-level truth standard was applied across all anatomical contexts and the study was not powered for diagnostic accuracy inference, this pattern is reported descriptively and may reflect protocol-related factors, including acquisition-time differences and the fixed scan order, rather than diagnostic superiority of either tracer.

### Sensitivity and Subgroup Analyses

The sensitivity analysis (n = 20) produced consistent directionality with the primary malignant set (Table 3). Adding the two histology-negative or indeterminate index target lesions, paired median SUVmax difference was −5.50 (95% CI −6.95 to −2.90; p = 3.9 × 10⁻⁴), Passing–Bablok slope 1.188 (95% CI 1.067–1.696), raw Bland–Altman bias −6.46 (95% LoA −23.07 to +10.15), and log-Bland–Altman GMR 0.73 (95% LoA 0.40× to 1.32×) (Supplementary Table S5). The wider limits of agreement and larger slope estimate in the sensitivity set reflect inclusion of the two nonmalignant target lesions, where histopathology did not confirm prostate cancer tumor biology at the sampled target site. In the pelvic lymph node subgroup of the primary malignant set, paired SUVmax difference remained significant (two-sided exact Wilcoxon signed-rank p = 0.0039); the difference in the prostate/prostatectomy-bed or locoregional recurrence subgroup was directionally consistent but did not reach significance (exact Wilcoxon p = 0.063) and was underpowered for formal inference given n = 6 (Supplementary Figure S3).

## Discussion

In this preliminary within-patient comparison, [¹⁸F]FC303 produced lower measured normal-organ SUVmean and lower malignant index target lesion SUVmax than [⁶⁸Ga]Ga-PSMA-11, while tumor-to-liver and tumor-to-mediastinum ratios showed no statistically significant paired difference. Paired malignant-lesion SUVmax values were strongly rank-correlated, but Passing–Bablok regression and log-scale Bland–Altman analysis demonstrated systematic and proportional differences across the measurement range. [¹⁸F]FC303 therefore yielded lower measured uptake in the prespecified physiologic reference organs rather than higher lesion SUV under the tested clinical protocols. Because urinary bladder and ureteral activity were not quantified and all analyzed index targets were pelvic, prostate, prostate-bed, or locoregional lesions, this lower physiologic-organ uptake should not be equated with a lower urinary-tract background or a generalized pelvic conspicuity advantage. Absolute SUV values were not interchangeable with those from [⁶⁸Ga]Ga-PSMA-11.

A distinctive feature of this study is the granularity of truth-standard reporting. Because all participants were drawn from the parent FC303-3-2 Phase 3 trial, each index target lesion was prospectively designated, anatomically matched, and histopathologically sampled, with local pathologists blinded to PET findings. We distinguished two analysis sets based on the specificity of verification at the analyzed anatomical site: a primary malignant set (n = 18, lesion-level or target-region verified malignant at the analyzed site) and a sensitivity set (n = 20, adding two histology-negative or indeterminate target lesions). The two histology-negative lesions in the sensitivity set—atypical atrophic acini suspicious for radiation therapy effect, and fibroadipose tissue with nerve ganglion—may reflect nonmalignant PSMA-avid uptake, treatment-related atypia, or spatial mismatch between the PET focus and the sampled tissue. In particular, the pelvic nodal target that showed high uptake on both tracers but histology of fibroadipose tissue with nerve ganglion is best interpreted separately as a possible PSMA-avid ganglion pitfall or spatial sampling mismatch rather than as an ordinary histology-negative lesion. Whether they represent true false-positive tracer uptake or spatial mismatch between the PET-detected focus and the histology-sampled region cannot be fully distinguished retrospectively; the former would reflect tracer specificity, whereas the latter would reflect a sampling limitation. Both mechanisms are recognized in the PSMA PET literature and argue against inferring tracer-level accuracy from lesion-level classification in a small, heterogeneous cohort. Separating these sets allows tumor-specific uptake inference to be clearly decoupled from broader target-lesion behavior.

These findings must be interpreted within non-harmonized acquisition protocols. [¹⁸F]FC303 was imaged at approximately 105 min post-injection, whereas [⁶⁸Ga]Ga-PSMA-11 was imaged at 60 min; administered activities differed two-fold (370 vs 185 MBq); and the scan order was fixed. These differences reflect current clinical practice constrained by gallium-68’s short physical half-life and by the favorable delayed-imaging pharmacokinetics achievable with fluorine-18, but they are central interpretive constraints, not peripheral limitations. The observed lower [¹⁸F]FC303 normal-organ uptake may reflect a combination of intrinsic tracer pharmacokinetics, delayed clearance at the later imaging timepoint, and count-statistics differences. Matched multi-timepoint studies will be required to disentangle these contributions; we therefore frame the present findings as protocol-conditional observations. Because all [⁶⁸Ga]Ga-PSMA-11 scans preceded [¹⁸F]FC303 and treatment-state granularity (paired PSA, ongoing ADT/ARPI exposure) at the second scan was incomplete, therapy-related temporal modulation of PSMA expression cannot be excluded, and the lower measured [¹⁸F]FC303 SUVmax should be interpreted as a protocol-conditional measurement difference rather than evidence of intrinsically lower tumor binding.

The Passing–Bablok slope of 1.13 (95% CI 1.04–1.45) in the primary malignant set is an important quantitative finding. Because the 95% CI excludes unity, the relationship between the two tracers is consistent with proportional differences across the measurement range: the absolute difference between tracers grows with increasing uptake, rather than being a constant additive offset. The sensitivity analysis (n = 20) produced a compatible slope estimate (1.19; 95% CI excluding 1.0). Combined with a log-scale Bland–Altman geometric mean ratio of 0.75 and 95% limits of agreement of 0.42× to 1.35× on the ratio scale, these results indicate that direct transfer of [⁶⁸Ga]Ga-PSMA-11-derived SUV thresholds to [¹⁸F]FC303 should not be assumed without tracer-specific calibration. This has conceptual relevance for clinical scenarios in which absolute SUV thresholds guide decision-making, including response assessment during systemic therapy and eligibility determination for ^177^Lu-PSMA radioligand therapy, where liver-relative uptake criteria are used in VISION/PSMAfore-type selection frameworks and absolute SUV thresholds were incorporated in TheraP-like selection approaches; however, the present cohort included only one patient with mCRPC and no analyzed bone lesions, and therefore does not establish [¹⁸F]FC303-specific thresholds for mCRPC or bone-dominant disease.^23–26^ The present data do not themselves establish calibrated thresholds; they identify the need for dedicated calibration studies.

Paired tumor-to-liver and tumor-to-mediastinum ratios in the primary malignant-lesion set did not differ significantly between tracers, with 95% CIs for the paired log ratios crossing unity. Because the study was not powered for equivalence or non-inferiority, these findings should be interpreted as absence of detected difference rather than evidence of equivalent contrast. The directionally higher T/L ratio with [¹⁸F]FC303 is compatible with the lower physiologic liver uptake observed in the normal-organ analyses.

Several additional limitations warrant emphasis. Formal urinary bladder and ureteral SUV quantification was not pre-specified and was not feasible retrospectively; because all analyzed index target lesions were locoregional (pelvic lymph node, prostate/prostatectomy-bed, or locoregional recurrence) targets, urinary tract activity is a particularly relevant unmeasured background compartment for prospective study. For pelvic lymph node specimens, pathology was frequently reported at the nodal basin level rather than by individual PET-imaged node, so verification should be interpreted as lesion-level or target-region histopathology. Reader independence was limited: tracer identity could not be fully masked because biodistribution patterns inherently reveal the tracer, and pre-consensus independent reads were not retained, precluding inter-reader agreement analysis. Paired PSA and detailed on-treatment status (ADT duration, ARPI exposure) at the time of [¹⁸F]FC303 PET/CT were not systematically captured, although no patient experienced documented systemic therapy change between examinations. The single-center design, retrospective nature, single-index-lesion approach, and limited sample size (n = 18 primary; n = 20 sensitivity) constrain generalizability. Scanner-level reconstruction parameters (algorithm, time-of-flight and point-spread-function modeling, iterations/subsets, and voxel size) were not fully documented for retrospective tabulation, and the sample was too small for a scanner-stratified sensitivity analysis; same-system within-patient pairing partially mitigates, but does not eliminate, this potential source of measurement variability. Although same-system pairing within each patient was confirmed from acquisition metadata, the specific distribution of patients across the two scanner models (Discovery MI and Omni Legend) and scanner-level reconstruction metadata were not uniformly retrievable for model-stratified tabulation. Administered ligand mass and molar activity at injection were likewise not systematically retrievable from the retrospective radiopharmaceutical records, so potential mass-dose effects on quantitative uptake could not be evaluated. Although histopathologic sampling followed the parent-trial window of 12 h to 28 days after [¹⁸F]FC303 PET/CT, exact histopathology dates were not uniformly retrievable, so the interval from [⁶⁸Ga]Ga-PSMA-11 PET/CT to histopathology could not be quantified per patient; given the fixed [⁶⁸Ga]-first order (median [⁶⁸Ga]-to-[¹⁸F]FC303 interval, 29.5 days; range, 19–60), it was necessarily longer than the corresponding [¹⁸F]FC303-to-pathology interval. The cohort included no analyzed bone lesions; findings should not be extrapolated to bone-metastatic disease or radioligand-therapy-eligible mCRPC populations, where tracer comparison is clinically most consequential.

Notwithstanding these limitations, this study provides, to our knowledge, the first paired within-patient quantitative reference data for [¹⁸F]FC303 against [⁶⁸Ga]Ga-PSMA-11, with same-system paired imaging within each patient and lesion-level or target-region histopathologic verification for all analyzed index target lesions. Prospective multicenter studies with harmonized or randomized acquisition protocols, systematic urinary-tract assessment, independent inter-reader analysis, and tracer-specific threshold calibration will be needed to define the clinical role of [¹⁸F]FC303 within PSMA-directed imaging workflows. Standardization frameworks such as E-PSMA and PROMISE (V2) offer a structured basis for such harmonization.^27, 28^

## Conclusions

In this preliminary within-patient comparison, [¹⁸F]FC303 PET/CT acquired under a delayed clinical acquisition protocol showed lower measured SUVmean in prespecified physiologic reference organs and lower malignant target-region SUVmax than [⁶⁸Ga]Ga-PSMA-11, while tumor-to-liver and tumor-to-mediastinum ratios showed no statistically significant paired difference in the primary malignant analysis set (n = 18). Because uptake time, scan order, administered activity, urinary-tract background, and treatment-state were not controlled, these findings should not be interpreted as tracer-intrinsic superiority or diagnostic-performance equivalence. Absolute SUV values were not interchangeable between tracers, with systematic and proportional differences across the measurement range (Passing–Bablok slope 1.13, 95% CI 1.04–1.45); direct transfer of [⁶⁸Ga]Ga-PSMA-11 SUV thresholds to [¹⁸F]FC303 should not be assumed. Consistent directionality was observed in the n = 20 sensitivity analysis. These data define protocol-conditional cross-tracer SUV constraints for [¹⁸F]FC303 rather than diagnostic superiority or transferable [⁶⁸Ga]Ga-PSMA-11–derived thresholds.

## Declarations

### Funding

FutureChem Co., Ltd. funded the parent FC303-3-2 Phase 3 clinical trial (NCT05936658) from which the study population was derived and provided radiopharmaceutical manufacturing and quality control support. Apart from the declared coauthor contributions of FutureChem employees to radiopharmaceutical preparation and quality control, FutureChem had no role in the design of the present retrospective analysis, image interpretation, statistical analysis, interpretation of the comparative findings, or the decision to submit the manuscript for publication.

### Competing Interests

W.J.L. and C.P. are employees of FutureChem Co., Ltd. R.K. is an employee of Inocras. W.-A.K., S.P., T.-S.K., and J.Y.J. declare no competing interests. Image interpretation and statistical analysis for this retrospective comparison were performed by academic investigators at the National Cancer Center of Korea, who were blinded to clinical/pathologic information and to the paired alternate-tracer result. Raw imaging and clinical data access was restricted to the academic corresponding authors. The decision to submit the manuscript for publication was made solely by the academic authors.

### Ethics Approval and Consent to Participate

The study protocol was approved by the Institutional Review Board of the National Cancer Center of Korea (IRB No. NCC2023-0090). The requirement for written informed consent was waived given the retrospective analysis of previously consented trial participants. All procedures complied with the Declaration of Helsinki.^18^

### Consent for Publication

Not applicable (no individually identifiable data are presented).

### Availability of Data and Materials

The datasets generated and analyzed during the current study are not publicly available due to institutional policies and parent-trial data-governance restrictions, but de-identified paired SUV data are available from the corresponding authors on reasonable request and subject to a data-use agreement.

### Authors’ Contributions

W.-A.K. and S.P. contributed equally as co-first authors; T.-S.K. and J.Y.J. contributed equally as corresponding authors. Conceptualization: W.-A.K., S.P., T.-S.K., J.Y.J. Data acquisition and image analysis: S.P., T.-S.K. Radiopharmaceutical preparation and quality control: W.J.L., C.P. Quantitative data processing and statistical programming: R.K. Statistical analysis and figure preparation: W.-A.K., S.P. Manuscript drafting: W.-A.K., S.P. Critical revision: all authors. All authors reviewed and approved the final manuscript.

## Acknowledgements

The authors thank the clinical and imaging staff at the National Cancer Center of Korea and the study coordinators of the FC303-3-2 Phase 3 trial.

## Notes

### Competing Interest Statement

FutureChem Co., Ltd. funded the parent FC303-3-2 Phase 3 clinical trial from which the study population was derived and provided radiopharmaceutical manufacturing and quality control support. W.J.L. and C.P. are employees of FutureChem Co., Ltd. R.K. is an employee of Inocras. The remaining authors declare no competing interests.

### Author Declarations

The study protocol was approved by the Institutional Review Board of the National Cancer Center of Korea (IRB No. NCC2023-0090). The requirement for written informed consent was waived given the retrospective analysis of previously consented trial participants. All procedures complied with the Declaration of Helsinki.

